# Electrocardiogram-based deep learning enables scalable screening of transthyretin amyloid cardiomyopathy

**DOI:** 10.1101/2024.09.30.24314651

**Authors:** Philip M. Croon, Lovedeep S. Dhingra, Bruno Batinica, Ryan B. Choi, Evangelos K. Oikonomou, Sumukh V. Shankar, Veer Sangha, Robert M.A. van der Boon, Michelle Michels, Maarten van Ettinger, Peter-Paul Zwetsloot, Sergio Teruya, Cesia Gallegos Kattan, Edward J. Miller, Navid Noory, Oscar M. Westin, Sie K. Fensman, Steen Hvitfeldt Poulsen, Avneet Singh, Sudarshan Balla, Anouk van Achten, Charalambos Vlachopoulos, Alexios S. Antonopoulos, Nico Bruining, Julian D. Gillmore, Mathew S. Maurer, Frederick L. Ruberg, Marianna Fontana, Rohan Khera

## Abstract

Transthyretin amyloid cardiomyopathy (ATTR-CM) is a treatable but underrecognized cause of heart failure, with diagnosis often delayed until advanced disease manifests. This gap is amplified in underserved populations at increased risk for ATTR-CM where access to specialist evaluation and advanced cardiac imaging is limited. Electrocardiograms (ECGs) are ubiquitous and often obtained years before ATTR-CM diagnosis in affected individuals, but conventional interpretation lacks the sensitivity and specificity needed for a practical screening tool. Here, we develop an artificial intelligence model that identifies ATTR-CM directly from widely available images of 12-lead ECGs. The model achieved an area under the receiver operating characteristic curve (AUROC) of 0.87 (95% confidence interval [CI], 0.82–0.91), with performance maintained across patients with echocardiographic features mimicking ATTR-CM. Performance was consistent and generalizable across 8 multinational validation cohorts with a wide range of prevalences across the US and Europe. Prospective deployment across three screening cohorts spanning older Black and Hispanic adults with heart failure and individuals with prior carpal tunnel syndrome surgery demonstrated clinical applicability with increased risk and plausible screening settings. These findings establish ECG imaging as a scalable entry point for ATTR-CM detection, enabling targeted referral for confirmatory testing and earlier initiation of disease-modifying therapy.

---

Transthyretin amyloid cardiomyopathy (ATTR-CM) is a progressive infiltrative cardiomyopathy caused by myocardial deposition of misfolded transthyretin fibrils.^1^ Once considered rare, ATTR-CM is now recognized as substantially underdiagnosed, with prevalence estimates ranging from 14-28% in heart failure with preserved ejection fraction and up to 14% in severe aortic stenosis.^2–5^ With the advent of disease-modifying therapies that slow progression and improve survival, early identification of patients with ATTR-CM has become a clinical priority.^6–8^ However, diagnoses still occur after the onset of advanced heart failure, when therapeutic benefit is attenuated, contributing to a poor median survival.^9–11^ This persistent gap between therapeutic opportunity and timely diagnosis underscores the urgent need for scalable, proactive screening strategies to identify ATTR-CM at its earliest detectable stages.

Current diagnostic approaches are not suited to population-wide screening, as they primarily rely on specialized, expensive, or invasive modalities, such as cardiac amyloid radionuclide imaging (CARI) or tissue biopsy. Electrocardiography (ECG), in contrast, is inexpensive, ubiquitous, routinely obtained across clinical settings, and is often available years before ATTR-CM is formally diagnosed.^12^ While cardiac amyloidosis can leave recognizable ECG signatures, conventional ECG measures associated with ATTR-CM lack the sufficient sensitivity and specificity required for screening.^13–16^ Artificial intelligence-enabled ECG (AI-ECG) offers a potential solution by detecting complex, disease-specific patterns, with early studies that reported promising preliminary results for the identification of ATTR-CM.^17–19^ However, prior models have been derived from highly selected cohorts with high prevalence of disease and have not been validated across heterogeneous clinical environments and in low prevalent cohorts, rendering them unsuitable as screening tools.^20^ Moreover, prior models rely upon raw signal data, requiring access to raw waveforms that typically are unavailable at the point of care, where ECGs are more commonly accessible as images.^21^ Given these shortcomings, there is an urgent need for an easily deployable, workflow-compatible approach that is validated across a wide range of realistic conditions.

In this study, we developed and validated a clinically deployable AI-ECG solution for detecting ATTR-CM from widely available ECG images. Following internal validation within a large US health system, we conducted a rigorous evaluation across eight distinct cohorts with a wide range of prevalences spanning the US and Europe. To support deployment across heterogeneous clinical environments, we built a locally deployable, format-agnostic inference pipeline accepting both routine ECG images and raw 12-lead signals, which enabled consistent retrospective validation across five multinational cohorts with diverse formats and vendors. Critically, we established the model’s clinical utility in three prospective, biologically enriched screening cohorts that reflect clinically representative populations in which ATTR-CM is actively sought but frequently underrecognized.

## Results

### Study Population

To develop the AI-ECG model, we used a convolutional neural network based on the EfficientNet-B3 architecture, leveraging 12-lead ECGs drawn from individuals receiving care within the Yale New Haven Health System(YNHHS), a large integrated health system encompassing five academic and community hospitals. To enable label-efficient model development, the network was initialized with weights from a pretrained EfficientNet-B3 model trained in 78,288 individuals with a self-supervised contrastive learning framework to recognize individual patient-specific patterns in ECGs, independent of their clinical interpretation.^22^ This pretrained model was fine-tuned using 28,174 12-lead ECGs from 11,291 unique patients. ECGs recorded between August 2015 and June 2023 were used for model development **(Supplementary Table 1)**. Cases were identified through chart review based on positive CARI or initiation of an approved transthyretin stabilizer. For each ECG from a person with ATTR-CM, we identified 20 ECGs from age- and sex-matched controls without ATTR-CM. The mean age was 80.4D±D8.7 years, 2,329 (20.6%) were women, and 9,385 (83.1%) individuals self-identified as White, 1,084 (9.6%) as Black, and 464 (4.1%) as Hispanic. The population had a high comorbidity burden, with 10,164 (90.0%) individuals having hypertension, 4,238 (37.5%) with diabetes, and 6,072 (53.8%) with heart failure **(Supplementary Table 1)**. Overall, 293 individuals (2.6%) with a total of 1,365 ECGs (4.8%) had confirmed cardiac amyloidosis.

Internal validation was performed in a temporally distinct held-out test cohort spanning July 2023 to July 2025, comprising 20,222 unique patients **(Table 1)**. Importantly, no patients in the held-out test cohort were present in the development cohort. The mean age was 67.4D±D16.5 years, and 9,753 (48.2%) were female. Of all individuals, 14,385 (71.1%) self-reported as White, 2,718 (13.4%) as Black, and 1,868 (9.2%) as Hispanic. History of hypertension was present in 16,244 (80.3%), diabetes in 6,540 (32.3%), and heart failure in 7,546 (37.3%) individuals. In total, 65 individuals (0.3%) had a confirmed cardiac amyloidosis diagnosis.

**Table 1:** Baseline characteristics of the internal validation cohorts.

|  | All patients<br>Total | All patients<br>Amyloid+ | All patients<br>Amyloid- | Mimics<br>Total | Mimics<br>Amyloid+ | Mimics<br>Amyloid- | CARI<br>Total | CARI<br>Amyloid+ | CARI<br>Amyloid- |
| --- | --- | --- | --- | --- | --- | --- | --- | --- | --- |
| **N patients** | 20222 | 65 | 20157 | 1900 | 65 | 1835 | 442 | 64 | 378 |
| **Total ECGs** | 61377 | 747 | 60630 | 5696 | 747 | 4949 | 5073 | 736 | 4337 |
| **Age** | 67.4 ± 16.5 | 81.1 ± 9.5 | 67.4 ± 16.5 | 74.0 ± 14.3 | 81.1 ± 9.5 | 73.7 ± 14.3 | 74.3 ± 11.6 | 81.0 ± 9.6 | 73.1 ± 11.5 |
| **Sex Female** | 9753.0 (48.2%) | 8.0 (12.3%) | 9745.0 (48.3%) | 786.0 (41.4%) | 8.0 (12.3%) | 778.0<br>(42.4%) | 182.0<br>(41.2%) | 8.0 (12.5%) | 174.0<br>(46.0%) |
| **Race/Ethnicity** |  |  |  |  |  |  |  |  |  |
| **White** | 14385 (71.1%) | 49 (75.4%) | 14336 (71.1%) | 1341 (70.6%) | 49 (75.4%) | 1292 (70.4%) | 322 (72.9%) | 48 (75.0%) | 274 (72.5%) |
| **Black** | 2718 (13.4%) | 10 (15.4%) | 2708 (13.4%) | 339 (17.8%) | 10 (15.4%) | 329 (17.9%) | 76 (17.2%) | 10 (15.6%) | 66 (17.5%) |
| **Hispanic** | 1868 (9.2%) | 2 (3.1%) | 1866 (9.3%) | 135 (7.1%) | 2 (3.1%) | 133 (7.2%) | 22 (5.0%) | 2 (3.1%) | 20 (5.3%) |
| **Missing** | 641 (3.2%) | 4 (6.2%) | 637 (3.2%) | 55 (2.9%) | 4 (6.2%) | 51 (2.8%) | 14 (3.2%) | 4 (6.2%) | 10 (2.6%) |
| **Asian** | 387 (1.9%) |  | 387 (1.9%) | 18 (0.9%) |  | 18 (1.0%) | 2 (0.5%) |  | 2 (0.5%) |
| **Others** | 223 (1.1%) |  | 223 (1.1%) | 12 (0.6%) |  | 12 (0.7%) | 6 (1.4%) |  | 6 (1.6%) |
| **Amyloidosis** | 65.0 (0.3%) | 65.0 (100.0%) | 0.0 (0.0%) | 65.0 (3.4%) | 65.0<br>(100.0%) | 0.0 (0.0%) | 64.0 (14.5%) | 64.0 (100.0%) | 0.0 (0.0%) |
| **Heart Failure** | 7546 (37.3%) | 38 (58.5%) | 7508 (37.2%) | 1067 (56.2%) | 38 (58.5%) | 1029 (56.1%) | 263 (59.5%) | 38 (59.4%) | 225 (59.5%) |
| **Acute Myocardial**<br>**Infarction** | 1277 (6.3%) | 4 (6.2%) | 1273 (6.3%) | 173 (9.1%) | 4 (6.2%) | 169 (9.2%) | 31 (7.0%) | 3 (4.7%) | 28 (7.4%) |
| **Ischemic Heart**<br>**Disease** | 5583 (27.6%) | 19 (29.2%) | 5564 (27.6%) | 669 (35.2%) | 19 (29.2%) | 650 (35.4%) | 123 (27.8%) | 18 (28.1%) | 105 (27.8%) |
| **Stroke** | 3561 (17.6%) | 10 (15.4%) | 3551 (17.6%) | 444 (23.4%) | 10 (15.4%) | 434 (23.7%) | 76 (17.2%) | 10 (15.6%) | 66 (17.5%) |
| **Hypertension** | 16244 (80.3%) | 60 (92.3%) | 16184 (80.3%) | 1779 (93.6%) | 60 (92.3%) | 1719 (93.7%) | 405 (91.6%) | 59 (92.2%) | 346 (91.5%) |
| **Diabetes** | 6540 (32.3%) | 15 (23.1%) | 6525 (32.4%) | 774 (40.7%) | 15 (23.1%) | 759 (41.4%) | 169 (38.2%) | 15 (23.4%) | 154 (40.7%) |

### Internal Validation of AI-ECG for the Detection of ATTR-CM

The AI-ECG model successfully identified cardiac amyloidosis across the overall test cohort and multiple clinically relevant subgroups **(Figure 2)**. In the full test set, the AI-ECG score yielded an area under the receiver operating characteristic curve (AUROC) of 0.87 (95% confidence interval [CI]: 0.82–0.91), with a sensitivity of 0.65 and specificity of 0.88 at the optimal threshold determined by the Youden index **(Figure 3)**. Model performance was consistent across demographic subgroups and was not affected by excluding individuals with AL amyloidosis **(Supplementary Figure 1)**.

**Figure 1:**
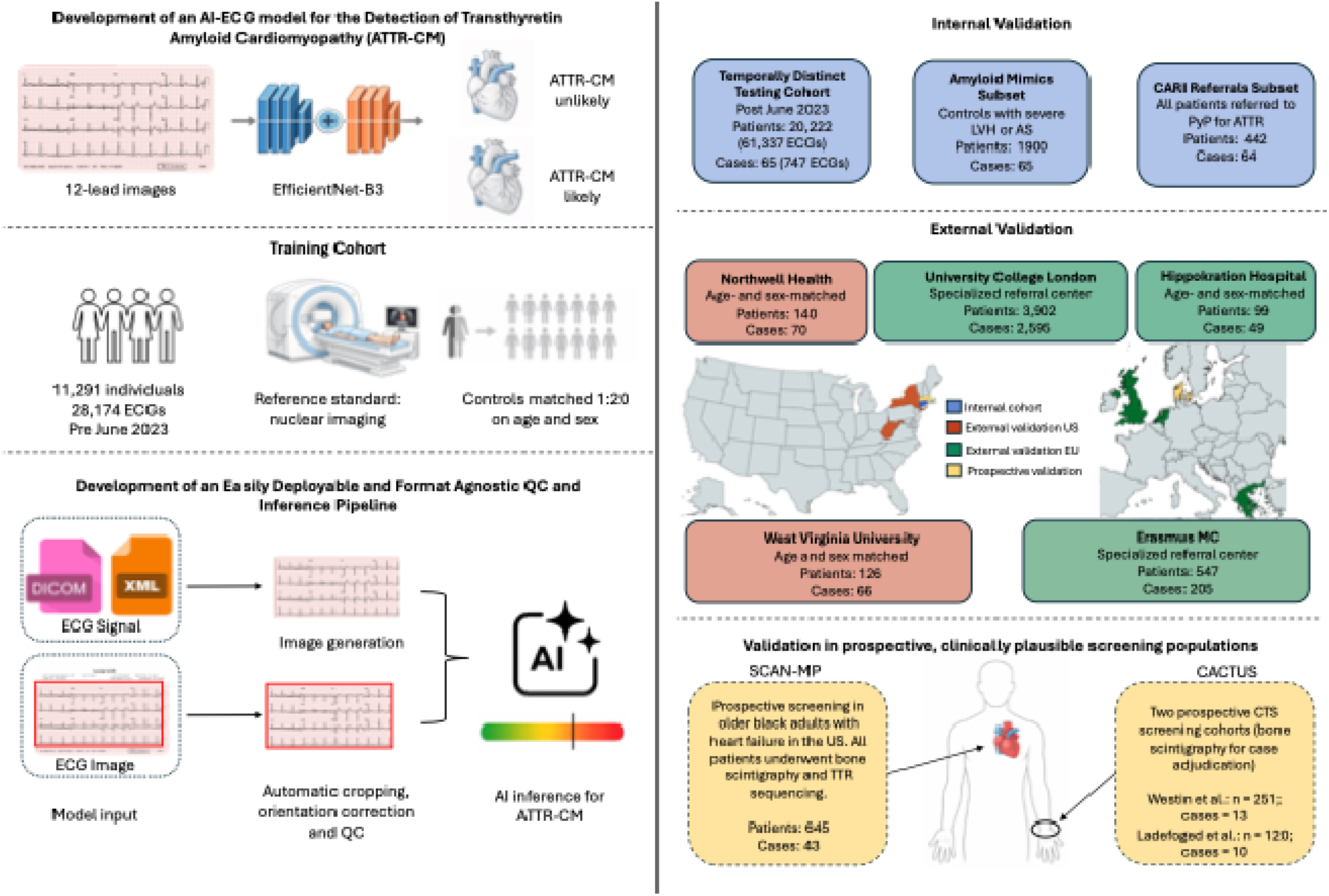
Development, validation, and real-world deployment of an AI-enabled electrocardiography pipeline for detection of transthyretin cardiac amyloidosis Overview of the development, internal validation, and multinational external validation of an artificial intelligence–enabled electrocardiography (AI-ECG) framework for detection of transthyretin amyloid cardiomyopathy (ATTR-CM). Left panel illustrates model development using 12-lead ECG images and an EfficientNet-B3 convolutional neural network, trained in a temporally split internal cohort from Yale New Haven Health System with case–control age- and sex-matching and adjudication of amyloid status by nuclear imaging. A format-agnostic quality control and inference pipeline enables standardized processing of ECG data derived from raw signals or heterogeneous ECG image formats through automated image generation, cropping, orientation correction, and quality control prior to AI inference. Right panel summarizes validation across internal test subsets, multinational external retrospective cohorts in the United States and Europe, and prospective clinically plausible screening populations, including individuals with heart failure and individuals with prior carpal tunnel syndrome surgery. The pipeline outputs a continuous AI-ECG risk score indicating likelihood of ATTR-CM, supporting scalable deployment across diverse clinical settings. **Abbreviations:** AI, artificial intelligence; ECG, electrocardiogram; AI-ECG, artificial intelligence–enabled electrocardiography; ATTR, transthyretin amyloidosis; ATTR-CM, transthyretin amyloid cardiomyopathy; CNN, convolutional neural network; QC, quality control; LVH, left ventricular hypertrophy; AS, aortic stenosis; CARI, cardiac amyloid radionuclide imaging; CTS, carpal tunnel syndrome; HF, heart failure; UCL, University College London.

**Figure 2:**
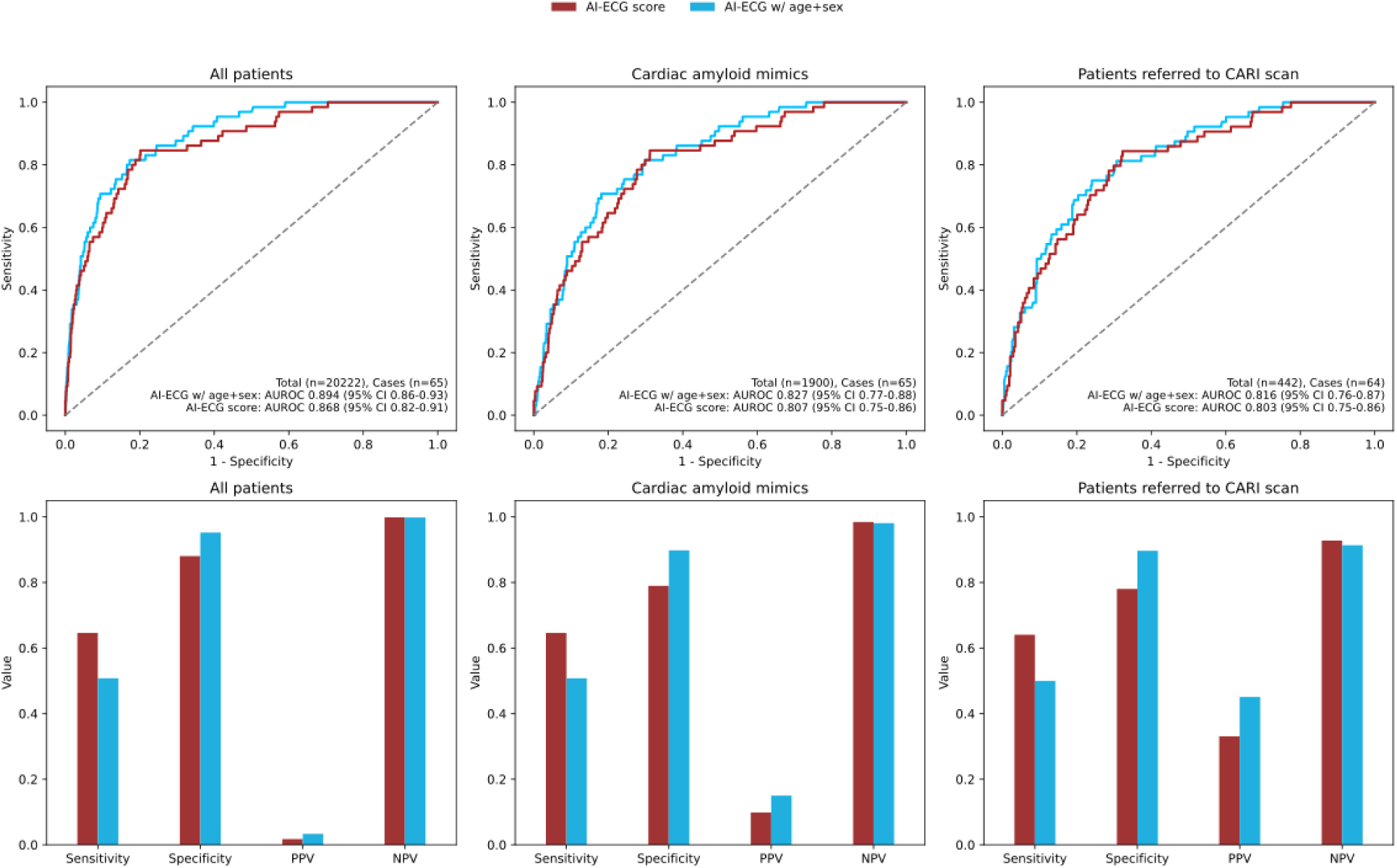
Discriminative Performance of the AI-ECG Model for Transthyretin Cardiac Amyloidosis Across Internal Subgroups Forest plot showing the AUROC with 95% confidence intervals of the AI-ECG model in the internal validation cohort. We present the AI-ECG score (red) and the ensemble model, which includes age and sex (blue). Subgroups include all patients, those with cardiac amyloid mimics (aortic stenosis, left ventricular hypertrophy, or transthyretin cardiac amyloidosis), and those referred for CARI. **Abbreviations:** AUROC, area under the receiver operating characteristic curve; PPV, positive predictive value; NPV, negative predictive value; AS, aortic stenosis; LVH, left ventricular hypertrophy; ATTR-CM, transthyretin cardiac amyloidosis; CARI, cardiac amyloid radionuclide imaging.

**Figure 3:**
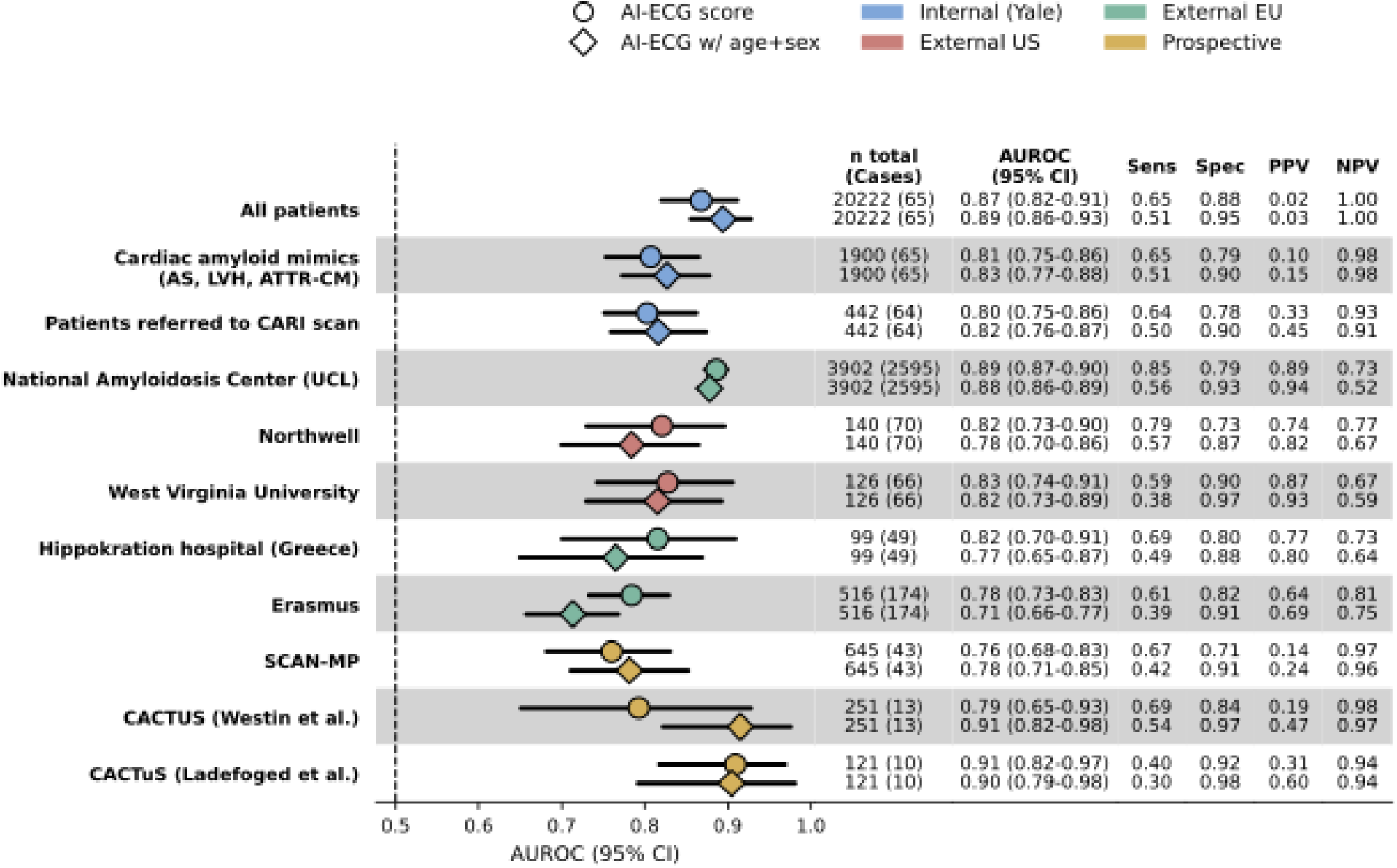
Performance of AI-ECG for detection of transthyretin cardiac amyloidosis across internal, external, and prospective cohorts. Forest plot showing discriminative performance of the AI-ECG score and an ensemble model incorporating age and sex (XGBoost) for detection of transthyretin cardiac amyloidosis (ATTR-CM) across internal, external, and prospective validation cohorts. Points indicate the area under the receiver operating characteristic curve (AUROC), with horizontal lines representing 95% confidence intervals. Cohorts are grouped by study design and geography, including internal validation in the Yale test cohort and clinically relevant subgroups, external retrospective validation in United States and European cohorts, and prospective validation in clinically plausible screening populations. Sensitivity, specificity, positive predictive value (PPV), and negative predictive value (NPV) are reported at a prespecified operating threshold. Cardiac amyloid mimics include patients with severe aortic stenosis (AS) and severe left ventricular hypertrophy (LVH). **Abbreviations:** AI-ECG, artificial intelligence–enhanced electrocardiography; AUROC, area under the receiver operating characteristic curve; CI, confidence interval; Sens, sensitivity; Spec, specificity; PPV, positive predictive value; NPV, negative predictive value; ATTR-CM, transthyretin cardiac amyloidosis; CARI, cardiac amyloid radionuclide imaging; LVH, left ventricular hypertrophy; AS, aortic stenosis.

Next, we performed additional external validation in two clinically relevant and high-comorbidity subgroups, starting with one comprising patients with clinical mimics of ATTR-CM, including those with aortic stenosis, left ventricular hypertrophy, or previously confirmed ATTR-CM. In this cohort, the AUROC for the AI-ECG score was 0.81 (95% CI: 0.75–0.86), with a sensitivity of 0.65 and specificity of 0.79 at the Youden-optimized threshold **(Figures 2, 3)**. Next, we deployed the model to the ECGs of all patients referred for CARI at YNHHS. Here, the AI-ECG score yielded an AUROC of 0.80 (95% CI: 0.75–0.86), with a sensitivity of 0.64 and specificity of 0.78 **(Figures 2, 3)**. Model performance across a range of operating thresholds, including corresponding sensitivity, specificity, positive predictive value (PPV), and negative predictive value (NPV), is summarized in **Supplementary Table 2**.

### Performance of AI-ECG in multinational external cohorts

External validation was performed across five independent cohorts spanning the US and Europe, comprising data from the National Amyloidosis Centre (NAC) in the United Kingdom and four age–and sex–matched case–control cohorts from Northwell Health, West Virginia University, Hippokration Hospital in Greece, and Erasmus MC in the Netherlands **(Table 2**, **Figure 2)**.

**Table 2:**
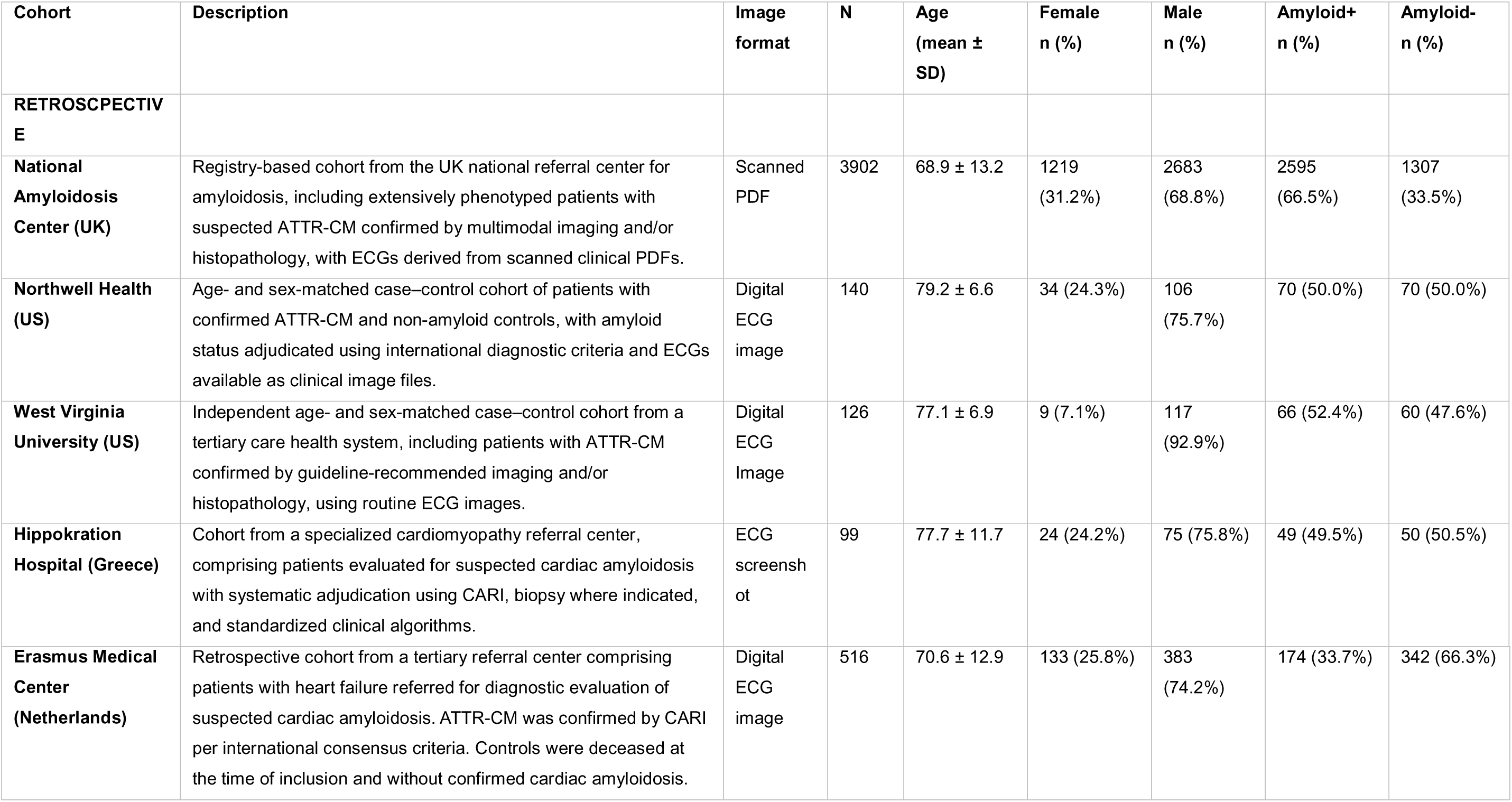
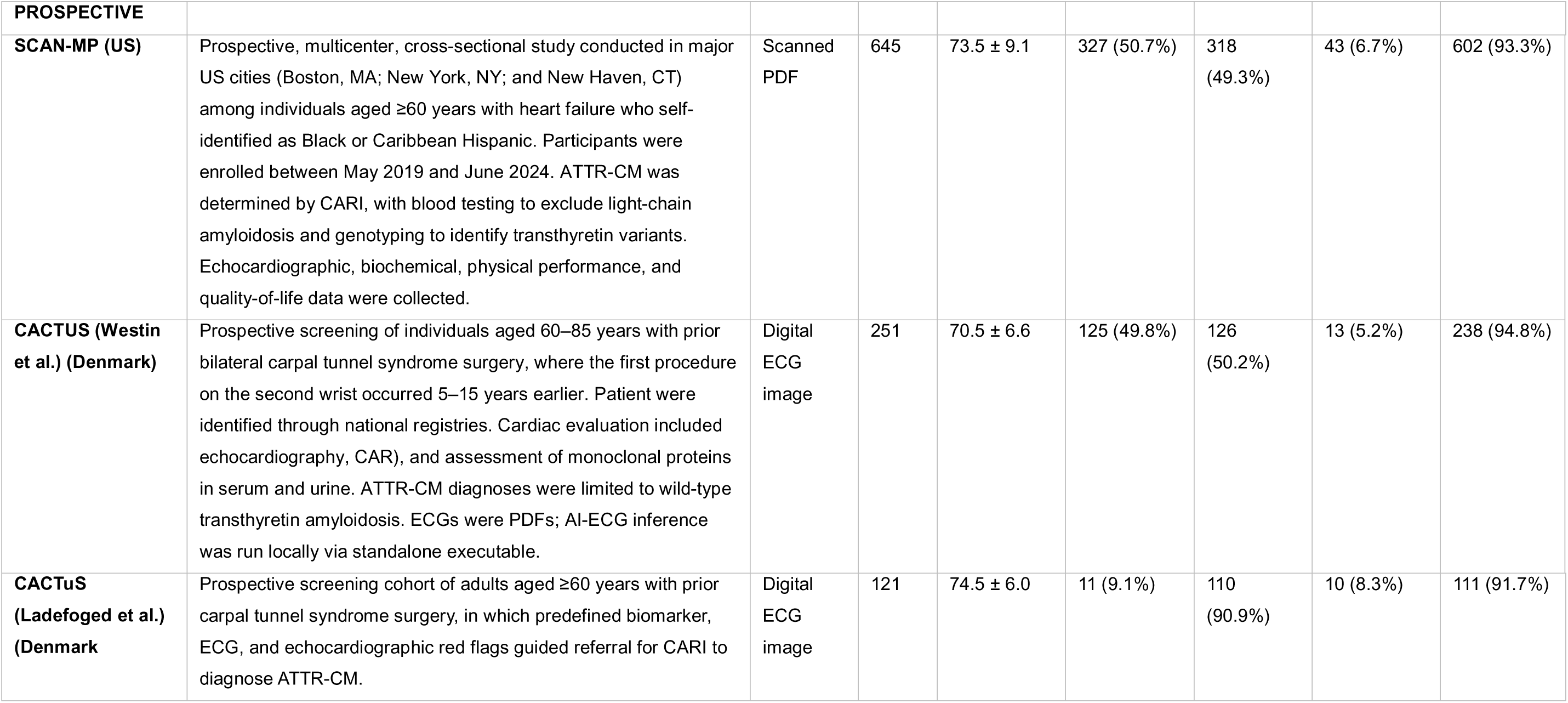
Description and baseline variables of the validation cohorts.

| Cohort | Description | Image format | N | Age (mean ± SD) | Female n (%) | Male n (%) | Amyloid+ n (%) | Amyloid- n (%) |
| --- | --- | --- | --- | --- | --- | --- | --- | --- |
| **RETROSPECTIVE** |  |  |  |  |  |  |  |  |
| **National Amyloidosis Center (UK)** | Registry-based cohort from the UK national referral center for amyloidosis, including extensively phenotyped patients with suspected ATTR-CM confirmed by multimodal imaging and/or histopathology, with ECGs derived from scanned clinical PDFs. | Scanned PDF | 3902 | 68.9 ± 13.2 | 1219 (31.2%) | 2683 (68.8%) | 2595 (66.5%) | 1307 (33.5%) |
| **Northwell Health (US)** | Age- and sex-matched case-control cohort of patients with confirmed ATTR-CM and non-amyloid controls, with amyloid status adjudicated using international diagnostic criteria and ECGs available as clinical image files. | Digital ECG image | 140 | 79.2 ± 6.6 | 34 (24.3%) | 106 (75.7%) | 70 (50.0%) | 70 (50.0%) |
| **West Virginia University (US)** | Independent age- and sex-matched case-control cohort from a tertiary care health system, including patients with ATTR-CM confirmed by guideline-recommended imaging and/or histopathology, using routine ECG images. | Digital ECG Image | 126 | 77.1 ± 6.9 | 9 (7.1%) | 117 (92.9%) | 66 (52.4%) | 60 (47.6%) |
| **Hippokration Hospital (Greece)** | Cohort from a specialized cardiomyopathy referral center, comprising patients evaluated for suspected cardiac amyloidosis with systematic adjudication using CARI, biopsy where indicated, and standardized clinical algorithms. | ECG screenshot | 99 | 77.7 ± 11.7 | 24 (24.2%) | 75 (75.8%) | 49 (49.5%) | 50 (50.5%) |
| **Erasmus Medical Center (Netherlands)** | Retrospective cohort from a tertiary referral center comprising patients with heart failure referred for diagnostic evaluation of suspected cardiac amyloidosis. ATTR-CM was confirmed by CARI per international consensus criteria. Controls were deceased at the time of inclusion and without confirmed cardiac amyloidosis. | Digital ECG image | 516 | 70.6 ± 12.9 | 133 (25.8%) | 383 (74.2%) | 174 (33.7%) | 342 (66.3%) |
| **PROSPECTIVE** |  |  |  |  |  |  |  |  |
| **SCAN-MP (US)** | Prospective, multicenter, cross-sectional study conducted in major US cities (Boston, MA; New York, NY; and New Haven, CT) among individuals aged $\geq 60$ years with heart failure who self-identified as Black or Caribbean Hispanic. Participants were enrolled between May 2019 and June 2024. ATTR-CM was determined by CARI, with blood testing to exclude light-chain amyloidosis and genotyping to identify transthyretin variants. Echocardiographic, biochemical, physical performance, and quality-of-life data were collected. | Scanned PDF | 645 | $73.5 \pm 9.1$ | 327 (50.7%) | 318 (49.3%) | 43 (6.7%) | 602 (93.3%) |
| **CACTUS (Westin et al.) (Denmark)** | Prospective screening of individuals aged 60–85 years with prior bilateral carpal tunnel syndrome surgery, where the first procedure on the second wrist occurred 5–15 years earlier. Patient were identified through national registries. Cardiac evaluation included echocardiography, CAR), and assessment of monoclonal proteins in serum and urine. ATTR-CM diagnoses were limited to wild-type transthyretin amyloidosis. ECGs were PDFs; AI-ECG inference was run locally via standalone executable. | Digital ECG image | 251 | $70.5 \pm 6.6$ | 125 (49.8%) | 126 (50.2%) | 13 (5.2%) | 238 (94.8%) |
| **CACTuS (Ladefoged et al.) (Denmark)** | Prospective screening cohort of adults aged $\geq 60$ years with prior carpal tunnel syndrome surgery, in which predefined biomarker, ECG, and echocardiographic red flags guided referral for CARI to diagnose ATTR-CM. | Digital ECG image | 121 | $74.5 \pm 6.0$ | 11 (9.1%) | 110 (90.9%) | 10 (8.3%) | 111 (91.7%) |

We first validated the AI-ECG model in 3,902 individuals referred to the NAC, one of the largest amyloidosis registries worldwide, with detailed phenotyping based on multimodal imaging, histopathology, and genetic testing. The mean age in this cohort was 68.9 ± 13.2 years, 1,219 participants (31.2%) were female, and all had amyloidosis, including 2,595 (66.5%) with confirmed cardiac involvement. This cohort enabled validation of the model in a highly specialized setting using digital ECG PDFs acquired as part of routine clinical care. The AI-ECG model achieved an AUROC of 0.89 (95% CI: 0.87–0.90). At the Youden-optimized threshold, the model achieved a sensitivity of 85.2% and specificity of 78.7%.

The detailed phenotyping at the NAC, including amyloid subtype classification by histopathology and genetic testing, enabled subtype-specific sensitivity analyses comparing patients with versus without cardiac involvement within each amyloid subtype. The model discriminated cardiac from non-cardiac involvement among patients with ATTR amyloidosis (n = 1,913; AUROC 0.94 [95% CI: 0.92–0.95]) and in patients with AL amyloidosis (n = 1,301; AUROC 0.89 [95% CI: 0.87–0.91]) **(Supplementary Table 3)**. Among ATTR subtypes, performance for detecting cardiac involvement was consistent in patients with wild-type ATTR (n = 1,353; AUROC 0.92 [95% CI: 0.89–0.95]) and individuals with a hereditary variant (n = 560; AUROC 0.93 [95% CI: 0.91–0.95]).

We further validated the model in four independent clinical populations, using a case–control design. In the cohorts from Hippokration Hospital in Athens, Northwell Health, and West Virginia University, ECGs were downloaded directly from the local electronic health record as site-native clinical images and analyzed using our local pipeline without reformatting or harmonization. In the Erasmus MC cohort, raw ECG signals were used to generate images for model evaluation.

At Northwell, AI-ECG achieved an AUROC of 0.82 (95% CI, 0.73–0.90) in 140 age- and sex-matched patients (mean age 79.2 ± 6.6 years; 34 [24%] women). Similar discrimination was observed at West Virginia University (n = 126; mean age 77.1 ± 6.9 years; 9 [7%] women), with an AUROC of 0.83 (95% CI, 0.74–0.91). At a specialized cardiomyopathy referral center at Hippokration Hospital (n = 99; mean age 77.7 ± 11.7 years; 24 [24%] women), with ATTR-CM adjudicated using standardized CARI protocols, AI-ECG achieved an AUROC of 0.82 (95% CI: 0.73–0.91). In the Erasmus Medical cohort (n = 516; 174 ATTR-CM cases; mean age 70.6 ± 12.9 years; 133 [25.8%] women), the model achieved an AUROC of 0.78 (95% CI: 0.73–0.83), with a sensitivity of 61.5%, specificity of 82.5%, and a PPV of 64.1% at the Youden-optimized threshold **(Figure 2)**. In a sensitivity analysis excluding patients with AL amyloidosis (n = 36), model performance was preserved (AUROC 0.79 [95% CI: 0.73–0.84]).

### Performance of AI-ECG for Detecting ATTR-CM in Clinically Relevant Prospective Cohort Studies

We next evaluated the AI-ECG model in three independent prospective screening cohorts designed to detect ATTR-CM in at risk populations, in which participants underwent systematic screening with CARI. These included the Screening for Cardiac Amyloidosis with Nuclear imaging in Minority Populations (SCAN-MP) study, which enrolled older individuals with heart failure, restricted to those of Black or Hispanic ancestry in whom the pV142I transthyretin variant is common, as well as two CACTUS studies that enrolled individuals with a history of bilateral carpal tunnel syndrome (CTS) surgery. All ECGs were analyzed as scanned PDF images acquired as part of these studies.

In the SCAN-MP cohort, which included 645 older Black and Hispanic participants with heart failure, 43 (6.7%) had a positive CARI scan **(Table 2)**. The median age was 73 years (interquartile range [IQR], 66–80). In this cohort, the AI-ECG model achieved an AUROC of 0.76 (95% CI, 0.68–0.83), with a sensitivity of 67% and specificity of 71% at the Youden-optimized threshold, yielding a PPV of 14% and an NPV of 97% **(Figure 3)**.

We then assessed the performance of the model in two independent carpal-tunnel-syndrome screening cohorts conducted at separate centers with non-overlapping participants. In the first CACTUS cohort (Westin et al.), which enrolled 251 adults with prior bilateral CTS and a median age of 70.4 years, 13 participants (5.2%) were diagnosed with ATTR-CM **(Table 2**, **Figure 3)**.^23^ Here, the AI-ECG model demonstrated an AUROC of 0.79 (95% CI, 0.65–0.93), a sensitivity of 0.69, and a specificity of 0.84.

In a second, independent CACTUS cohort (Ladefoged et al.; n = 121; 10 ATTR-CM cases) performance was excellent, demonstrating an AUROC of 0.91 (95% CI, 0.82–0.97), a sensitivity of 0.40, and a specificity of 0.92 for AI-ECG **(Table 2**, **Figure 3)**.

### Age- and sex-augmented ensemble model

Recognizing that ATTR-CM prevalence varies substantially by age and sex, we integrated these variables with convolutional neural network (CNN) output in a secondary XGBoost ensemble model to determine whether simple demographic information could improve discrimination. This ensemble model provided modest incremental improvement over the standalone CNN across non-age- and sex-matched cohorts. In internal validation, the ensemble model achieved an AUROC of 0.89 (95% CI: 0.86–0.93), compared with 0.87 for the CNN alone, and similar gains were observed in the cardiac amyloid mimic subgroup (ensemble AUROC 0.83 vs. CNN 0.81) and in patients referred for CARI (ensemble AUROC 0.82 vs. CNN 0.80). In the prospective screening cohorts, the ensemble model yielded AUROCs of 0.78 (95% CI: 0.71–0.85) in SCAN-MP, 0.92 (95% CI: 0.82–0.98) in CACTUS (Westin), and 0.90 (95% CI: 0.79–0.98) in CACTuS (Ladefoged), compared with CNN AUROCs of 0.76, 0.79, and 0.91, respectively. In SCAN-MP, the ensemble model increased the PPV from 14% to 24% while maintaining an NPV of 96%. Full ensemble results across all cohorts are reported in **Table 2** and **Supplementary Table 4**.

### Comparison with an echocardiography-based clinical score

To assess whether an AI-ECG model based solely on routine ECG images could match the performance of a screening approach that requires less readily available echocardiography, we benchmarked the CNN model against an established echocardiography-based ATTR-CM risk score (Davies et al.) in 18,088 patients with available echocardiographic data (89.4% of the full internal test cohort).^24^ Across all patients, the AI-ECG model achieved an AUROC of 0.88 (95% CI, 0.82–0.94), comparable to the echocardiographic score (AUROC 0.87; 95% CI, 0.82–0.92) **(Figure 4; Supplementary Table 5 and Supplementary Figure 2)**.

**Figure 4:**
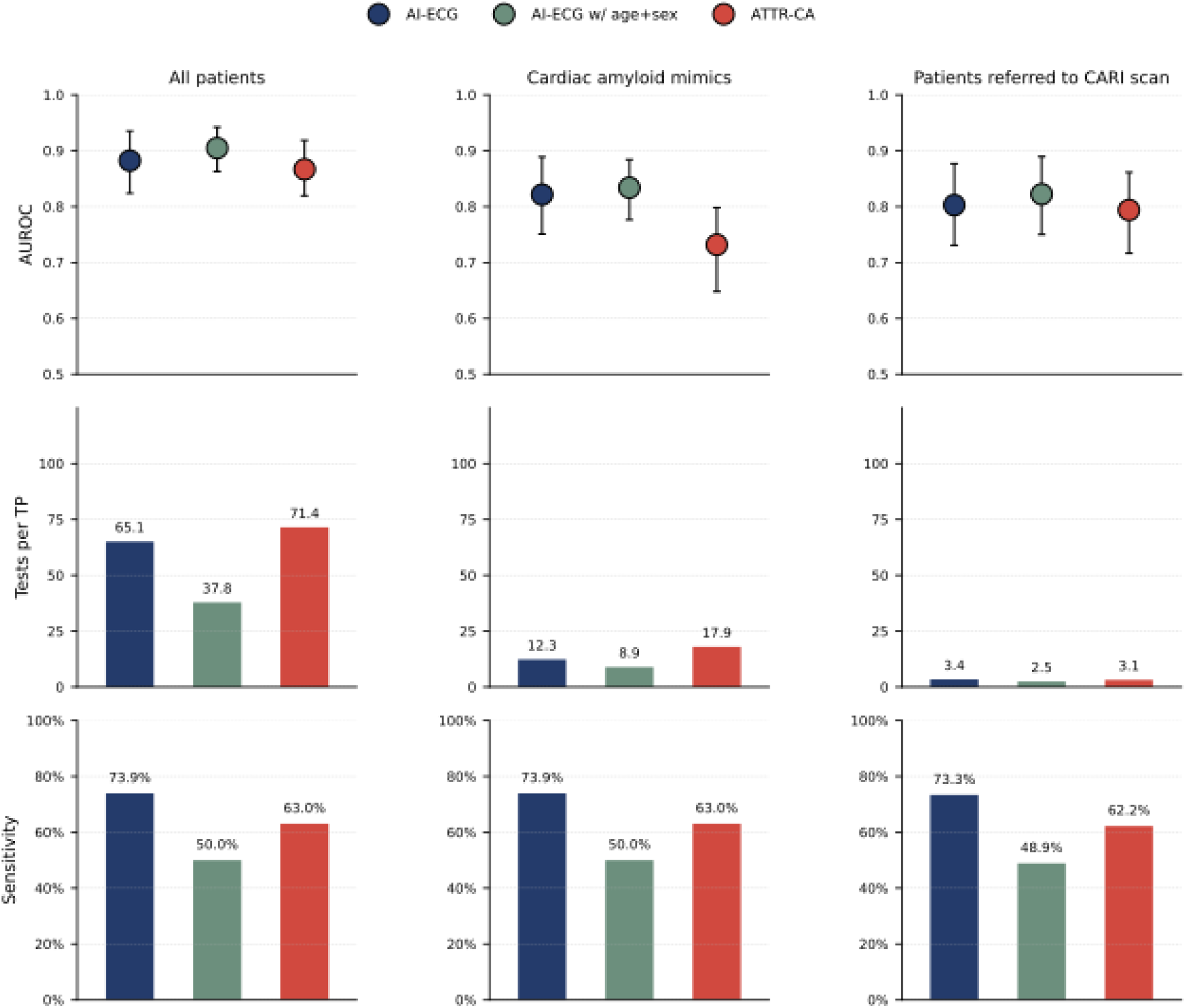
Comparative discriminative performance, clinical yield, and sensitivity of AI-ECG, clinical, and echocardiographic risk models across clinically relevant screening populations Comparison of an artificial intelligence–enhanced electrocardiogram (AI-ECG) model, a clinical model incorporating the CNN predictions and age and sex (CNN + age/sex), and an echocardiography-based ATTR-CM risk score across three clinically relevant cohorts: all patients, patients with cardiac amyloid mimics, and patients referred for cardiac amyloid radionuclide imaging. The cohort is a subset of the total cohort including everyone with an echocardiogram. The top row shows the area under the receiver operating characteristic curve (AUROC) with 95% confidence intervals. Middle row shows clinical yield expressed as the number of CARI scans required per true positive diagnosis (tests per true positive; 1/PPV) at a fixed threshold yielding a specificity of 85%. Bottom row shows sensitivity at the same fixed threshold. Cardiac amyloid mimics include patients with severe aortic stenosis and/or severe left ventricular hypertrophy. AI-ECG, artificial intelligence–enhanced electrocardiography; AUROC, area under the receiver operating characteristic curve; ATTR-CM, transthyretin amyloid cardiomyopathy; CI, confidence interval; CNN, convolutional neural network; PPV, positive predictive value; CARI, cardiac amyloid radionuclide imaging.

In the cardiac amyloid mimic cohort, the AI-ECG model yielded an AUROC of 0.82 (95% CI, 0.75–0.89), compared with 0.73 (95% CI, 0.65–0.80) for the echocardiographic score **(Figure 4; Supplementary Table 5)**. Similarly, in patients referred for CARI, the AI-ECG model achieved an AUROC of 0.80 (95% CI, 0.73–0.88) versus 0.79 (95% CI, 0.72–0.86) for the clinical score **(Figure 4)**. Clinical yield for the AI-ECG model was 3.4 tests per true-positive diagnosis, compared with 3.1 tests for the echocardiography-based score **(Figure 4; Supplementary Table 5)**.

### Model Activation Visualizations for Representative Examples

To qualitatively examine ECG regions contributing to model predictions, we generated gradient-weighted class activation map (Grad-CAM) visualizations for representative true-positive, true-negative, false-positive, and false-negative cases **(Supplementary Figure 3)**. In the representative true-positive example, activation was most prominent over leads showing visually apparent ECG abnormalities, including low voltages and QRS widening. In the illustrated false-positive example, activation involved similar ECG regions despite the absence of adjudicated ATTR-CM. By contrast, the true-negative and false-negative examples showed comparatively limited activation, consistent with less marked electrocardiographic abnormalities.

## Discussion

In this study, we develop and validate an accurate and scalable platform for the detection of ATTR-CM from images of routinely performed 12-lead ECG images or signals using a newly developed AI-ECG model. Across a temporally distinct internal test cohort and eight validation cohorts spanning the US and Europe the model maintained clinically relevant performance despite substantial heterogeneity in acquisition workflows, image formats, and patient populations. Performance was preserved in clinically complex settings where phenotypic overlap often complicates recognition and improved modestly with the addition of age and sex. These findings position format-agnostic AI-ECG, deployable on either routine ECG images or raw signal data, as a practical frontline triage strategy to support earlier diagnosis of ATTR-CM in real-world care.

These findings should be interpreted in the context of ongoing efforts to improve early detection of ATTR-CM using AI. Prior AI-based approaches have largely focused on echocardiography, where deep learning models applied to echocardiographic videos have demonstrated strong discrimination.^24–27^ However, their potential as frontline screening tools is constrained by the resources needed to obtain an echocardiogram. In contrast, the electrocardiogram is inexpensive, widely available, and routinely acquired across care settings, making it well-suited for scalable deployment. Prior studies have demonstrated the promise of AI-ECG for detecting cardiac amyloidosis. Grogan et al. showed that a signal-based AI-ECG model could identify cardiac amyloidosis, though validation was limited to the institution where the model was developed.^18,28^ Goto et al. subsequently reported external validation at two additional institutions.^17^ However, these early studies were based on internal or case-control matched cohorts from US centers and required access to raw ECG signal data, which is often unavailable at the point of care.

In the present study, we substantially extended this work by developing a format-agnostic AI-ECG model that operates on both routine ECG images and raw waveform signals, in the largest ATTR-CM cohort assembled to date and validating it across a large number of multinational cohorts with data from registries and studies in the US and Europe. This framework incorporated heterogeneous, locally obtained ECG formats across sites, including raw digital signals, vendor-specific image exports, and scanned images. The ECG format agnostic pipeline allowed for the deployment of the model locally by the participating site reducing the chance of developer bias.^29^ To our knowledge, this is the first AI-ECG model for transthyretin cardiac amyloidosis to be evaluated through local deployment across a broad range of realistic clinical settings, including a highly specialized national referral cohort with comprehensive phenotyping, multiple tertiary referral centers in Europe and the US, and three independent prospective screening cohorts. Together, these validation settings provide a rigorous test of model performance across diverse acquisition workflows, data standards, and intended-use populations.

From a clinical standpoint, the consistent performance observed in clinically complex populations with a wide range of prevalences highlights its potential utility at multiple points along the ATTR-CM care pathway, including settings where symptoms and phenotypes overlap and where multimorbidity often complicates referral decisions. First, as a pragmatic screening tool, AI-ECG demonstrated consistent performance across biologically informed prospective screening cohorts, namely in older Black and Hispanic individuals with heart failure and individuals with prior CTS surgery where ATTR-CM is prevalent.^30–32^ The comparison with an established echocardiography-based ATTR-CM risk score further supports this positioning. In the subset of patients with available echocardiographic data, AI-ECG achieved discrimination comparable to, and in some clinically relevant subgroups exceeding, that of the echocardiography-based score, despite relying solely on a routinely acquired ECG. This supports its use as an ECG-first triage strategy for referral to confirmatory imaging. Second, in clinically complex patients with substantial phenotypic overlap, including those with left ventricular hypertrophy or aortic stenosis in whom adjudicating cardiac involvement remains challenging, the model’s performance remained strong. Similarly, the model’s performance was excellent within a dedicated amyloidosis referral center, where it discriminated between cardiac and non-cardiac involvement in individuals with known amyloidosis. Third, prior studies demonstrating that AI-ECG scores track preclinical progression of ATTR-CM over time raise the possibility that AI-ECG may also serve as a biomarker of disease burden with potential utility for longitudinal monitoring.^12,33^ Rather than replacing definitive testing, AI-ECG may support a stepwise, resource-efficient care pathway that improves diagnostic yield, optimizes the use of confirmatory imaging, and expands access to timely diagnosis and disease-modifying therapy.^34^

Several limitations should be acknowledged. First, ATTR-CM remains a rare disease in the general population, and disease prevalence varies across cohorts, thereby inherently constraining the positive predictive value in unselected settings. However, rather than solely relying on artificially enriched case–control datasets, this study deliberately evaluated AI-ECG performance in clinically feasible prospective screening cohorts that reflect contemporary diagnostic practice, thereby providing more realistic estimates of clinical yield. Second, although external cohorts included ECGs acquired in heterogeneous formats, such as scanned paper ECGs and images exported from different vendors, careful validation would be required to ensure the reliability of predictions in novel formats. Third, despite rigorous case adjudication, ATTR-CM remains widely underrecognized in clinical practice. Some individuals labeled as controls may have had unrecognized or early-stage ATTR-CM, thereby attenuating performance. Finally, this study did not assess the downstream clinical impact of AI-ECG–guided screening on diagnostic workflows, treatment initiation, or outcomes, which will require prospective trials.

## Conclusion

AI-ECG enables scalable detection of ATTR-CM from images of routinely acquired 12-lead ECGs. Across referral and prospective screening cohorts, it showed clinically relevant performance under real-world acquisition conditions, including scanned PDFs from heterogeneous vendor formats and raw signal data. These findings support AI-ECG as a practical frontline triage tool for targeted confirmatory testing and earlier treatment initiation.

## Methods

This study leveraged multiple retrospective and prospective cohorts to develop and validate an AI-ECG model for the detection of ATTR-CM **(Figure 1)**. Model development used data from routine clinical care within a large integrated US health system. External validation was performed across five independent cohorts, including a national amyloidosis referral registry from the NAC in the United Kingdom, two US-based case–control cohorts, a specialized cardiomyopathy referral center in Greece, and a heart failure cohort from the Netherlands. In addition, model performance was evaluated in three prospective, biologically enriched screening studies. Our study follows the TRIPODD+DAI reporting guidelines **(Supplementary Material)**.^35^ The study protocol was approved by the Yale Institutional Review Board, which waived the requirement for informed consent because the analyses were conducted using existing clinical and research data.

### Data Sources and Study Population

The development and internal validation cohorts were derived from individuals receiving care within the YNHHS, a large integrated health system in the northeastern US comprising five academic and community hospitals and affiliated outpatient clinics.

We identified individuals who underwent a standard 12-lead ECG between August 2015 and June 2023 for model development. A temporally distinct cohort of ECGs recorded between July 2023 and July 2025 was reserved as a held-out internal test set for internal validation. Additionally, we created demographic subgroups based on age (<70 vs ≥70 years), sex, and self-identified race (Black or White) to identify potential model bias. Finally, we created a cohort excluding individuals with an ICD code for AL amyloidosis (E85.81) to conduct a sensitivity analysis aimed at understanding how AL amyloidosis affects the model’s performance. To prevent data leakage, no patients overlapped between the development and testing cohorts.

#### Case and Control Definitions

Cases of ATTR-CM were identified through electronic health record review and defined by either (i) a positive CARI scan or (ii) treatment with an approved transthyretin stabilizer. Across cohorts, CARI was performed in accordance with American Society of Nuclear Cardiology recommendations using single-photon emission–computed tomography, with study positivity adjudicated by the interpreting physician based on semiquantitative visual grading (grade ≥2).^36^ The earliest occurrence of either diagnostic criterion was designated as the diagnosis date, and all ECGs obtained within 12 months before or after this date were eligible for inclusion.

Controls were defined as individuals who did not meet the above criteria for ATTR-CM and who did not have relevant ICD codes for amyloidosis (E85.2, E85.82). ECGs of individuals with ATTR-CM were matched on a 1:20 ratio to ECGs from controls. Development data were split into training and validation at a 9:1 ratio. The number of ECGs per individual in the control group was restricted to a maximum of five to ensure balanced representation across patients and to limit the influence of individuals with frequent ECG acquisition.

### ECG Preprocessing and Image Generation

During model training, ECG waveform signals were converted to grayscale images using a custom Python-based plotting pipeline that simulates clinical variation in ECG appearance. This included variability in lead layout, the inclusion and positioning of rhythm strips, differences in grid density, line thickness, and font size or position. These preprocessing steps ensured that the model was trained on a diversity of ECG formats encountered in clinical workflows and vendor-specific formats. All images were plotted at a standard calibration of 10 mm/mV and uniformly downsampled to 300 × 300 pixels. To improve generalizability across heterogeneous ECG formats, images were randomly rotated by up to ±10 degrees during data augmentation. This methodology has been described in previous reports.^37,38^

### AI-ECG Model Architecture and Internal Testing

The model architecture and training procedure is described in detail in the Supplementary Methods. In short, the model architecture was a CNN based on the EfficientNet-B3 architecture. To facilitate label-efficient learning, the model was initialized with weights from a self-supervised pretraining task in which a separate cohort of ECGs was used to train the network to identify individual-specific ECG patterns irrespective of clinical interpretation.^22^ Model training was performed in two stages. In the initial stage, the final four layers were unfrozen and fine-tuned using a learning rate of 0.01 for two epochs. Subsequently, all layers were unfrozen and trained using a reduced learning rate of 5 × 10⁻D for six additional epochs. Training used the Adam optimizer with gradient clipping and a minibatch size of 64. Early stopping was applied based on validation loss, with training halted if no improvement was observed for three consecutive epochs. Importantly, no ECGs used during pretraining overlapped with those included in model development or validation.

### Internal validation across clinically relevant subgroups

To evaluate internal validity in clinically relevant and challenging settings, we assessed model performance within predefined subgroups in the held-out validation cohort. These subgroups included patients with cardiac amyloid mimics and all patients referred for CARI. Cardiac amyloid mimics were defined as the presence of left ventricular hypertrophy (LVH), severe aortic stenosis (AS), or confirmed ATTR-CM. LVH was defined as an interventricular septal thickness >14 mm on echocardiography or documentation of LVH in the echocardiography report. Severe aortic stenosis was defined according to standard echocardiographic criteria as the presence of a peak aortic jet velocity ≥4.0 m/s, a mean transvalvular gradient ≥40 mmHg, an aortic valve area <1.0 cm², or an indexed aortic valve area <0.6 cm²/m².^39–41^

### Development of the Locally Deployable Inference Pipeline

To support deployment across heterogeneous clinical environments, we developed a locally deployable ECG inference pipeline that can process both raw signal data and routine clinical ECG images. This framework was designed to enable standardized model inference within partner institutions without requiring manual image editing, centralized reformatting, or access to a uniform vendor-specific ECG format.

For signal-based ECGs, waveform data were extracted from XML or DICOM files and rendered using the same image-generation pipeline used during model training, without augmentation. For image-based ECG inputs, including scanned ECGs and PDF exports from electronic health records, we developed a dedicated preprocessing workflow based on a YOLOv8 object detection model trained to identify ECG tracings. This workflow automatically verifies the presence of a valid ECG, isolates the ECG region, removes surrounding borders and identifiable patient information, corrects image orientation, and generates standardized inputs for model inference. To facilitate use across sites, the full preprocessing and inference pipeline was packaged for both server-based deployment and local execution for Apple and Windows operating systems.

### External retrospective validation cohorts

We validated the performance of the AI-ECG model in five multinational cohorts, including cohorts derived from specialty referral centers **(Table 2)**.

First, we evaluated performance in the NAC, a uniquely informative referral cohort in which all participants had confirmed amyloidosis and cardiac involvement had been comprehensively adjudicated. Individuals in this cohort undergo comprehensive, standardized phenotyping for suspected amyloidosis, including multidisciplinary clinical evaluation, biomarker assessment, and advanced multimodality cardiac imaging. ATTR-CM diagnoses were adjudicated according to international consensus criteria using CARI, cardiovascular magnetic resonance, and, where indicated, histopathology. Control individuals without cardiac amyloidosis were drawn from the same referral population. AI-ECG inference was performed directly on scanned ECG image files using the standardized deployment pipeline.

Next, we utilized the local deployment pipeline to validate the model’s performance, starting with two independent US-based healthcare systems: Northwell Health and West Virginia University Health System. In both cohorts, retrospective age- and sex-matched case–control designs were used, with individuals with ATTR-CM matched to control patients without evidence of amyloidosis. ATTR-CM diagnoses were adjudicated according to international consensus criteria, primarily using CARI in conjunction with clinical assessment. ECGs were available as image files exported from the electronic health record, and AI-ECG inference was performed directly on these images using the standardized, format-agnostic deployment pipeline.

We further assessed model performance in a retrospective cohort derived from patients seeking care in the specialized cardiomyopathy referral center of the Hippokration Hospital in Greece. Individuals with ATTR-CM were age- and sex-matched to cardiomyopathy patients without amyloidosis drawn from the same referral population. ATTR-CM diagnoses were adjudicated in accordance with international guidelines, primarily using CARI, with biopsy performed when clinically indicated. AI-ECG inference was performed directly on ECG images obtained during routine clinical care.

Finally, we evaluated model performance in a retrospective cohort from Erasmus Medical Center (Rotterdam, the Netherlands), comprising patients who were referred for diagnostic evaluation of suspected cardiac amyloidosis. Cases with confirmed ATTR-CM were identified based on CARI and clinical assessment according to standard international consensus criteria. Control individuals without confirmed cardiac amyloidosis, including carriers of pathogenic *TTR* DNA variants without an established phenotype were drawn from the same referral population. ECG images were available from the electronic health record, and AI-ECG inference was performed using the standardized, format-agnostic deployment pipeline.

### Prospective validation in clinically important screening cohorts

We assessed the utility of AI-ECG in high-risk and biologically plausible subgroups using data from three prospective screening cohorts. The first prospective validation was conducted using data from the SCAN-MP study, a prospective multicenter cohort of older Black and Caribbean Hispanic adults with heart failure and an increased left ventricular wall thickness.^32,42^ ATTR caused by a *TTR* gene variant (*pV142I)* is common in this population, making it an important target for screening.^43,44^ In this cohort, ATTR-CM was confirmed by CARI and blood testing to exclude monoclonal proteins, and all individuals underwent *TTR* genotyping, allowing classification of both wild-type and variant ATTR-CM. ECGs were collected from multiple sites and available in different formats, including scanned printouts and digitally exported images from various machines.

The second prospective cohort was derived from the CACTUS study, a prospective cohort of adults aged 60–85 years with a history of bilateral CTS surgery. CTS is often caused by ATTR deposits in the transverse carpal tenosynovium and is strongly associated with ATTR-CM.^23,30,45^ Participants in this cohort were identified through national registries, and eligibility was determined by having undergone a second CTS surgical wrist procedure 5–15 years before enrollment. All participants underwent CARI and assessment of monoclonal proteins in serum and urine. ECG images collected at enrollment were included in the analysis.

The third prospective cohort was derived from a second CACTuS study.^46^ This screening cohort of adults aged ≥60 years included participants who were invited for screening after CTS surgery. Screening incorporated pre-specified “red flags,” including elevated cardiac biomarkers (N-terminal pro-B-type natriuretic peptide [NT-proBNP] and/or cardiac troponin), ECG patterns associated with ATTR-CM, echocardiographic left ventricular hypertrophy, and/or impaired longitudinal strain with apical sparing. Participants meeting any red-flag criterion underwent confirmatory CARI for ATTR-CM adjudication, and individuals without red flags were not routinely referred for CARI.

### Ensemble Modeling with Clinical Features

To assess whether incorporating basic demographic information improved discrimination, we trained a secondary ensemble model using gradient-boosted decision trees (XGBoost) that combined the continuous AI-ECG score with patient age and sex. The ensemble model was trained on the same development cohort as the convolutional neural network. Hyperparameters were optimized using grid search with three-fold cross-validation. This model was deployed and validated on all internal and external validation cohorts.

### Comparison with an echocardiographic clinical score

To benchmark the AI-ECG model against a risk score that requires echocardiography, we calculated the ATTR-CM risk score described by Davies et al. ^24^ We restricted this analysis to all patients in the held-out test set with both ECG and available echocardiographic data and evaluated performance across the all-patients cohort, the ATTR-CM mimic cohort, and all patients referred for CARI. The score was computed at the patient level using age, sex, hypertension status, and standard echocardiographic parameters, including left ventricular ejection fraction, posterior wall thickness, and relative wall thickness, defined as (interventricular septal thickness in diastole [IVSd] + left ventricular posterior wall thickness in diastole [LVPWd])/left ventricular internal diameter in diastole (LVIDd) and binarized using a threshold of 6. To enable a fair head-to-head comparison, analyses were restricted to a stable subset of patients with complete data and non-missing outputs for the AI-ECG model, the ensemble model, and the echocardiography-based risk score, ensuring that observed differences in performance reflected model characteristics rather than differences in cohort composition.

### Identification of predictive cues

To assess model interpretability, we applied gradient-weighted class activation mapping (Grad-CAM) to the final convolutional layers of the trained AI-ECG model, generating saliency maps that highlight ECG regions contributing most strongly to model predictions. Grad-CAM analyses were performed on representative true-positive, true-negative, false-positive, and false-negative ECGs.

### Statistical analysis

Categorical variables were summarized as counts and percentages (n [%]), and continuous variables as means with standard deviations (SD). To create a demographically balanced training cohort, we performed nearest-neighbor matching at the ECG level using the MatchIt package in R. ECGs from patients with confirmed amyloid cardiomyopathy were matched to control ECGs, with exact matching on sex and nearest-neighbor matching on age. Model performance was evaluated using the AUROC, with 95% confidence intervals estimated using bootstrapping with 500 random subsamples using 80% of the test set. For binary classification, the operating threshold was determined using the Youden index (maximizing the sum of sensitivity and specificity) in the validation set. At this threshold, sensitivity, specificity, PPV, and NPV were reported. The threshold was fixed based on the internal validation set; performance at this threshold in external and prospective cohorts reflects the operating characteristics of the model in those populations rather than a reoptimized threshold.

### Data and Code Sharing

The individual-level data underlying this study cannot be made publicly available due to patient privacy regulations and institutional data use agreements. Code used for statistical analysis will be made publicly available upon publication.

### Sources of funding

Dr Khera was supported by the National Institutes of Health (R01AG089981, R01HL167858, and K23HL153775) and the Doris Duke Charitable Foundation (2022060). Dr Oikonomou was supported by the National Heart, Lung, and Blood Institute of the National Institutes of Health (F32HL170592). Dr Maurer and Dr Ruberg were supported by the National Institutes of Health (R01HL139671) for the SCAN-MP cohort.

### Disclosures

RK reported receiving grants from the National Heart, Lung, and Blood Institute, National Institutes of Health, Doris Duke Charitable Foundation, Bristol Myers Squibb, Novo Nordisk, BridgeBio, Amgen, Ionis, and Blavatnik Foundation, being an academic cofounder of Ensight-AI and Evidence2Health, having patents 63/346,610, WO2023230345A1, US20220336048A1, 63/484,426, 63/508,315, 63/580,137, 63/606,203, 63/619,241, and 63/562,335 pending, and serving as associate editor of JAMA outside the submitted work. OW reported receiving speaker fees and advisory board compensation from AstraZeneca and an independent research grant from Pfizer. No other disclosures were reported. EKO acknowledges research support from the American Heart Association (AHA; award no. 26CDA1612298), the Robert A. Winn Excellence in Clinical Trials Career Development Award, and the Wiesman Award for Excellence in Early-Career ATTR Research from Cornerstone Medical Education through Yale University. He is a co-inventor on patent applications (filed through Yale University) and granted patents licensed through the University of Oxford to Caristo Diagnostics Ltd. He is a co-founder of Evidence2Health LLC, and has served as a consultant to Caristo Diagnostics Ltd and Ensight-AI Inc. He has also received honoraria from Clinical Education Alliance and serves as an Associate Editor for the European Heart Journal. These affiliations and potential financial interests have been disclosed and are being managed in accordance with institutional policies. FLR reports research funding from NIH (R01HL139671, R01HL177670, and R01AG093132), TriNetX/AstraZeneca, Pfizer, Anumana, and BridgeBio, and personal fees from eMyosound and Cardiovascular Core Laboratories.

## Supplemental Materials

Supplementary Tables 1-5

Supplementary Figures 1-3

## Data Availability

An online version of the model is publicly available for research use at https://www.cards-lab.org/ecgvision-attrcm. This web application represents a prototype of the eventual application of the model, with instructions for required image standards and a version that demonstrates an automated image standardization pipeline.

## Abbreviations

ATTR-CM: transthyretin amyloid cardiomyopathy
YNHHS: Yale New Haven Health System
NAC: National Amyloidosis Center
CARI: cardiac amyloid radionuclide imaging
Grad-CAM: gradient-weighted class activation mapping
IVSd: interventricular septal thickness in diastole
LVPWd: left ventricular posterior wall thickness in diastole
LVIDd: left ventricular internal diameter in diastole

