## Supplementary materials for "Electrocardiogram-based deep learning enables scalable screening of transthyretin amyloid cardiomyopathy"

**Supplementary tables**

**Table 1: Baseline characteristics of the training dataset**

|  | Train Total | Train Amyloid+ | Train Amyloid- |
| --- | --- | --- | --- |
| Patients | 11291 | 293 | 10998 |
| Age | 80.4 ± 8.7 | 82.0 ± 7.6 | 80.4 ± 8.8 |
| Sex Female | 2329 (20.6%) | 58 (19.8%) | 2271 (20.6%) |
| Race/Ethnicity |  |  |  |
| White | 9385 (83.1%) | 204 (69.6%) | 9181 (83.5%) |
| Black | 1084 (9.6%) | 62 (21.2%) | 1022 (9.3%) |
| Hispanic | 464 (4.1%) | 16 (5.5%) | 448 (4.1%) |
| Missing | 213 (1.9%) | 9 (3.1%) | 204 (1.9%) |
| Asian | 100 (0.9%) | 1 (0.3%) | 99 (0.9%) |
| Others | 45 (0.4%) | 1 (0.3%) | 44 (0.4%) |
| Heart Failure | 6072 (53.8%) | 229 (78.2%) | 5843 (53.1%) |
| Acute Myocardial Infarction | 1405 (12.4%) | 41 (14.0%) | 1364 (12.4%) |
| Ischemic Heart Disease | 4696 (41.6%) | 115 (39.2%) | 4581 (41.7%) |
| Stroke | 2494 (22.1%) | 51 (17.4%) | 2443 (22.2%) |
| Hypertension | 10164 (90.0%) | 264 (90.1%) | 9900 (90.0%) |
| Diabetes | 4238 (37.5%) | 101 (34.5%) | 4137 (37.6%) |
| ATTR-CM | 293 (2.6%) | 293 (100.0%) | 0 (0.0%) |

**Table 2: Performance metrics of the CNN across different thresholds**

| **Cohort** | **Threshold** | **Threshold value** | **Sensitivity** | **Specificity** | **PPV** | **NPV** | **AUROC** | **AUROC 95% CI lower** | **AUROC 95% CI upper** | **Cases** | **Controls** |
| --- | --- | --- | --- | --- | --- | --- | --- | --- | --- | --- | --- |
| All | Sensitivity 80% | 0.129 | 0.65 | 0.88 | 0.02 | 1.0 | 0.868 | 0.82 | 0.912 | 65 | 20157 |
| All | Sensitivity 90% | 0.033 | 0.85 | 0.72 | 0.01 | 1.0 | 0.868 | 0.82 | 0.912 | 65 | 20157 |
| All | Specificity 85% | 0.246 | 0.51 | 0.94 | 0.03 | 1.0 | 0.868 | 0.82 | 0.912 | 65 | 20157 |
| All | Youden (Max J) | 0.129 | 0.65 | 0.88 | 0.02 | 1.0 | 0.868 | 0.82 | 0.912 | 65 | 20157 |
| LVH/AS/Amyloid (Mimics) | Sensitivity 80% | 0.129 | 0.65 | 0.79 | 0.1 | 0.98 | 0.807 | 0.753 | 0.865 | 65 | 1835 |
| LVH/AS/Amyloid (Mimics) | Sensitivity 90% | 0.033 | 0.85 | 0.6 | 0.07 | 0.99 | 0.807 | 0.753 | 0.865 | 65 | 1835 |
| LVH/AS/Amyloid (Mimics) | Specificity 85% | 0.246 | 0.51 | 0.87 | 0.12 | 0.98 | 0.807 | 0.753 | 0.865 | 65 | 1835 |
| LVH/AS/Amyloid (Mimics) | Youden (Max J) | 0.129 | 0.65 | 0.79 | 0.1 | 0.98 | 0.807 | 0.753 | 0.865 | 65 | 1835 |
| CARI | Sensitivity 80% | 0.129 | 0.64 | 0.78 | 0.33 | 0.93 | 0.803 | 0.751 | 0.862 | 64 | 378 |
| CARI | Sensitivity 90% | 0.033 | 0.84 | 0.58 | 0.26 | 0.96 | 0.803 | 0.751 | 0.862 | 64 | 378 |
| CARI | Specificity 85% | 0.246 | 0.5 | 0.87 | 0.4 | 0.91 | 0.803 | 0.751 | 0.862 | 64 | 378 |
| CARI | Youden (Max J) | 0.129 | 0.64 | 0.78 | 0.33 | 0.93 | 0.803 | 0.751 | 0.862 | 64 | 378 |

**Table 3: AI-ECG model performance across cardiac amyloidosis subtypes in the National Amyloidosis Centre (NAC) validation cohort.**

| Subgroup | n | Cases | Controls | AUROC (95% CI) | Sensitivity | Specificity | PPV | NPV |
| --- | --- | --- | --- | --- | --- | --- | --- | --- |
| Overall | 3,902 | 2,595 | 1,307 | 0.886 (0.873–0.898) | 85.2% | 78.7% | 88.8% | 72.8% |
| ATTR (all TTR) | 1,913 | 1,661 | 252 | 0.938 (0.923–0.952) | 88.2% | 85.7% | 97.6% | 52.4% |
| ATTRv | 560 | 399 | 161 | 0.931 (0.909–0.954) | 81.7% | 90.1% | 95.3% | 66.5% |
| ATTRwt | 1,353 | 1,262 | 91 | 0.923 (0.894–0.948) | 90.3% | 78.0% | 98.3% | 36.6% |
| AL | 1,301 | 932 | 369 | 0.889 (0.867–0.911) | 79.9% | 86.2% | 93.6% | 63.0% |

Performance statistics shown at the Youden index-optimal threshold. AUROC, area under the receiver operating characteristic curve; CI, confidence interval; PPV, positive predictive value; NPV, negative predictive value; ATTR, transthyretin amyloid cardiomyopathy; ATTRv, variant transthyretin amyloid cardiomyopathy; ATTRwt, wild-type transthyretin amyloid cardiomyopathy; AL, light-chain amyloidosis.

**Table 4: Performance metrics of the ensemble XGBoost model across different thresholds**

| **Cohort** | **Threshold** | **Threshold value** | **Sensitivity** | **Specificity** | **PPV** | **NPV** | **AUROC** | **AUROC 95% CI lower** | **AUROC 95% CI upper** | **Cases** | **Controls** |
| --- | --- | --- | --- | --- | --- | --- | --- | --- | --- | --- | --- |
| All | Sensitivity 80% | 0.004 | 0.71 | 0.9 | 0.02 | 1.0 | 0.894 | 0.855 | 0.928 | 65 | 20157 |
| All | Sensitivity 90% | 0.002 | 0.82 | 0.8 | 0.01 | 1.0 | 0.894 | 0.855 | 0.928 | 65 | 20157 |
| All | Specificity 85% | 0.012 | 0.51 | 0.95 | 0.03 | 1.0 | 0.894 | 0.855 | 0.928 | 65 | 20157 |
| All | Youden (Max J) | 0.012 | 0.51 | 0.95 | 0.03 | 1.0 | 0.894 | 0.855 | 0.928 | 65 | 20157 |
| CARI | Sensitivity 80% | 0.004 | 0.7 | 0.79 | 0.36 | 0.94 | 0.816 | 0.759 | 0.874 | 64 | 378 |
| CARI | Sensitivity 90% | 0.002 | 0.81 | 0.64 | 0.28 | 0.95 | 0.816 | 0.759 | 0.874 | 64 | 378 |
| CARI | Specificity 85% | 0.012 | 0.5 | 0.9 | 0.46 | 0.91 | 0.816 | 0.759 | 0.874 | 64 | 378 |
| CARI | Youden (Max J) | 0.012 | 0.5 | 0.9 | 0.45 | 0.91 | 0.816 | 0.759 | 0.874 | 64 | 378 |
| LVH/AS/Amyloid (Mimics) | Sensitivity 80% | 0.004 | 0.71 | 0.8 | 0.11 | 0.99 | 0.827 | 0.772 | 0.878 | 65 | 1835 |
| LVH/AS/Amyloid (Mimics) | Sensitivity 90% | 0.002 | 0.82 | 0.67 | 0.08 | 0.99 | 0.827 | 0.772 | 0.878 | 65 | 1835 |
| LVH/AS/Amyloid (Mimics) | Specificity 85% | 0.012 | 0.51 | 0.9 | 0.15 | 0.98 | 0.827 | 0.772 | 0.878 | 65 | 1835 |
| LVH/AS/Amyloid (Mimics) | Youden (Max J) | 0.012 | 0.51 | 0.9 | 0.15 | 0.98 | 0.827 | 0.772 | 0.878 | 65 | 1835 |

**Table 5: Comparative diagnostic performance of AI-ECG–based models and an echocardiography-derived clinical risk score**

| Cohort | Model | N | AUROC (95% CI) | Sensitivity | Specificity | PPV | NPV | Tests per TP | Threshold |
| --- | --- | --- | --- | --- | --- | --- | --- | --- | --- |
| All patients | AI-ECG score | 18088 | 0.88 (0.82-0.94) | 0.74 | 0.88 | 0.02 | 1.00 | 65.1 | 0.129 |
| All patients | XGBoost (CNN, Age/Sex) | 18088 | 0.90 (0.86-0.94) | 0.50 | 0.95 | 0.03 | 1.00 | 37.8 | 0.012 |
| All patients | ATTR-CM score | 18088 | 0.87 (0.82-0.92) | 0.63 | 0.89 | 0.01 | 1.00 | 71.4 | 6.000 |
| Cardiac amyloid mimics | AI-ECG score | 1803 | 0.82 (0.75-0.89) | 0.74 | 0.78 | 0.08 | 0.99 | 12.3 | 0.129 |
| Cardiac amyloid mimics | XGBoost (CNN, Age/Sex) | 1803 | 0.83 (0.78-0.88) | 0.50 | 0.90 | 0.11 | 0.99 | 8.9 | 0.012 |
| Cardiac amyloid mimics | ATTR-CM score | 1803 | 0.73 (0.65-0.80) | 0.63 | 0.72 | 0.06 | 0.99 | 17.9 | 6.000 |
| Patients referred to CARI | AI-ECG score | 351 | 0.80 (0.73-0.88) | 0.73 | 0.75 | 0.30 | 0.95 | 3.4 | 0.129 |
| Patients referred to CARI | XGBoost (CNN, Age/Sex) | 351 | 0.82 (0.75-0.89) | 0.49 | 0.90 | 0.41 | 0.92 | 2.5 | 0.012 |
| Patients referred to CARI | ATTR-CM score | 351 | 0.79 (0.72-0.86) | 0.62 | 0.80 | 0.32 | 0.94 | 3.1 | 6.000 |

Discriminative performance is shown for the AI-ECG score, an ensemble model combining the AI-ECG score with age and sex (XGBoost), and an echocardiography-based ATTR-CM clinical risk score across all patients with available echocardiograms, a cardiac amyloid mimic cohort, and patients referred for cardiac amyloid radionuclide imaging (CARI). Area under the receiver operating characteristic curve (AUROC) is reported with 95% confidence intervals. Sensitivity, specificity, positive predictive value (PPV), negative predictive value (NPV), and tests per true positive diagnosis are reported at the Youden-optimized operating thresholds for AI-ECG and XGBoost models, and at the prespecified threshold for the ATTR-CM score. Tests per true positive represent the inverse of PPV. Abbreviations: AI-ECG, artificial intelligence–enhanced electrocardiography; AUROC, area under the receiver operating characteristic curve; ATTR-CM, transthyretin amyloid cardiomyopathy; CI, confidence interval; NPV, negative predictive value; PPV, positive predictive value; CARI, cardiac amyloid radionuclide imaging.

**Supplementary Figures**

**Figure 1: Performance of the AI-ECG Amyloidosis Model Across Demographic Subgroups**

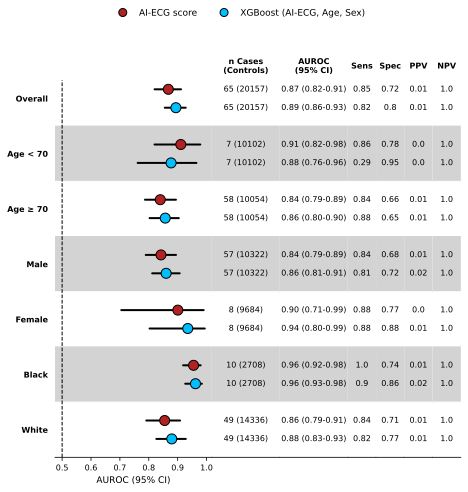

Abbreviations: AI-ECG, artificial intelligence–enhanced electrocardiography; AUROC, area under the receiver operating characteristic curve; CI, confidence interval; Sens, sensitivity; Spec, specificity; PPV, positive predictive value; NPV, negative predictive value.

**Supplementary Figure 2:** Receiver operating characteristic curves comparing AI-ECG, ensemble, and echocardiography-based risk models across clinically relevant cohorts

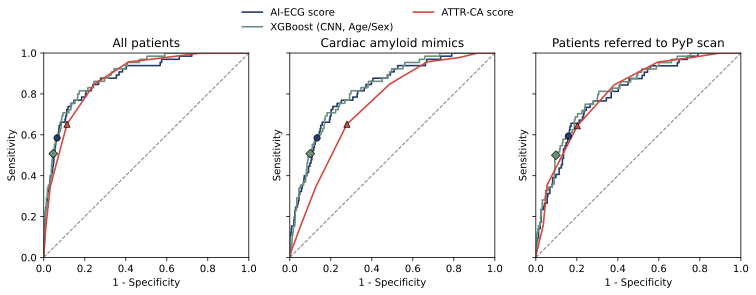

Receiver operating characteristic (ROC) curves comparing the discriminative performance of the AI-ECG model (blue), an ensemble model incorporating the AI-ECG score with age and sex using XGBoost (green), and an echocardiography-based ATTR-CM clinical risk score (red) across three cohorts: all patients with available echocardiograms (left), patients with cardiac amyloid mimics (middle), and patients referred for cardiac amyloid radionuclide imaging (CARI) (right). The dashed diagonal line represents performance no better than chance. Filled markers indicate the prespecified operating point for each model corresponding to the primary analysis threshold used for reporting sensitivity, specificity, and clinical yield. **Abbreviations:** AI-ECG, artificial intelligence–enabled electrocardiography; AUROC, area under the receiver operating characteristic curve; ATTR-CM, transthyretin amyloid cardiomyopathy; CARI, cardiac amyloid radionuclide imaging.

**Supplementary Figure 3** Model interpretability using gradient-weighted class activation mapping (Grad-CAM)

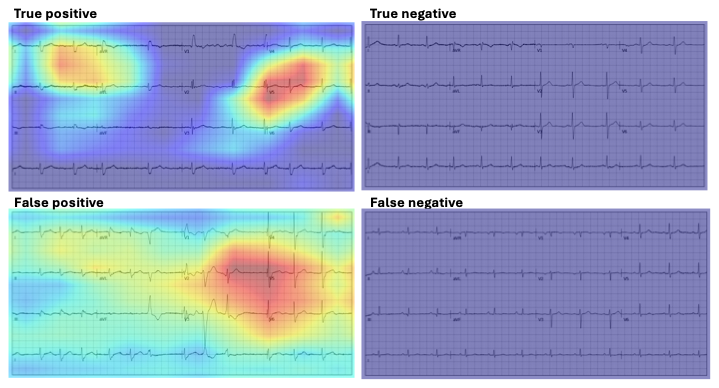

Gradient-weighted class activation maps (Grad-CAM) overlaid on representative 12-lead ECG images from true-positive, true-negative, false-positive, and false-negative predictions of the AI-ECG model. Warmer colors indicate regions contributing more strongly to the model’s predicted probability of transthyretin amyloid cardiomyopathy (ATTR-CM). Visualizations were generated from the final convolutional layer of the trained model and are intended to provide qualitative insight into model attention rather than mechanistic interpretation.
